# Development of a national osteopathic practice-based research network: the NCOR Research Network

**DOI:** 10.1101/2024.09.03.24312982

**Authors:** Jerry Draper-Rodi, Carol Fawkes, Daniel Bailey

## Abstract

**Objectives:** To describe the development of the NCOR Research Network, the first osteopathic Practice-Based Research Network (PBRN) in the UK, and provide data on its members’ characteristics, clinical practices, and patient demographics.

**Design:** Cross-sectional survey study.

**Setting:** Online survey of osteopaths practising in the United Kingdom.

**Participants:** 570 osteopaths registered with the General Osteopathic Council who consented to participate in the NCOR Research Network.

**Primary and secondary outcome measures:** Demographic characteristics of osteopaths, details of their clinical practice, patient demographics, common presenting complaints, treatment approaches, and attitudes towards evidence-based practice.

**Results:** The median age bracket of participants was 50-59 years, with 55% identifying as women. Participants had a median of 17 years of clinical experience. Most worked in private practice (71% as principals, 32% as associates), seeing 20-39 hours of patients per week. The majority (87%) regularly treated adults aged 65 or older. Low back pain was the most common complaint seen daily (56%). Spinal articulation/mobilization (79%) and soft tissue massage (78%) were the most frequently used techniques. Participants reported positive views towards evidence-based practice but cited lack of research skills and time as barriers to engagement.

**Conclusions:** The NCOR Research Network provides a foundation for future osteopathic research in the UK. While the sample was not fully representative of UK osteopaths, it offers insights into current osteopathic practice. The network aims to foster collaboration between clinicians and academics, potentially bridging the gap between research and practice in osteopathy.

**Protocol registration:** https://doi.org/10.17605/OSF.IO/HPWG4

**Article summary:** Strengths and limitations of this study:

- This study establishes the first osteopathic Practice-Based Research Network (PBRN) in the UK, providing a novel infrastructure for collaborative research in osteopathy.
- The survey was developed based on previous PBRN studies and existing UK osteopathic datasets, allowing for comparability of data across different research initiatives.
- A diverse recruitment strategy was employed, including various channels such as the regulatory body, professional organisations, and both face-to-face and online presentations, to reach a wide range of osteopaths.
- The sample in this study is not fully representative of the UK osteopathic profession when compared to the General Osteopathic Council registrant data, which may limit the generalisability of the findings.
- As the survey data were self-reported by osteopaths, the findings may be subject to recall or social desirability biases.

## Introduction

Osteopathy is a regulated profession in the United Kingdom (UK) and is an Allied Health Profession (AHP) in England. Osteopaths principally manage patients with persistent musculoskeletal (MSK) presentations, delivering packages of care using a variety of strategies including manual therapy, self-management, education, and reassurance [1]. Most osteopaths in the UK are self-employed and work alone [1]. In order to promote reflective learning and interaction of osteopaths with other professionals, the General Osteopathic Council (GOsC), the professional regulator, changed the Continuing Professional Development requirements in 2018 to include a mandatory objective activity to invite osteopaths to discuss cases with colleagues, collect patient feedback or data using Patient Reported Outcome Measures (PROMs), peer observation, or conduct a clinical audit [2]. Around the country, there are several regional societies to promote collaboration and shared learning opportunities for osteopaths, as working in isolation is seen as a potential risk for burnout and patient safety issues [3]. However, accessibility to these societies due to geographical dispersion may be limited [4]. Consequently, there is a need to develop an easily accessible network of osteopaths to support shared learning and participation in activities.

There seems to be a gap between the patient care delivered in osteopathic practices throughout the UK and the existing evidence. An umbrella review found promising evidence regarding osteopathic care for MSK disorders, but limited and inconclusive evidence for paediatric conditions, primary headache and irritable bowel syndrome [5]. However, non-MSK disorders are commonly treated by osteopaths in clinical practice, despite the lack of evidence for this approach [1]. Consequently, we need a better understanding of what happens in osteopathic clinical practice: we need more evidence, of better quality, about what osteopaths do in their clinics; and we need to assess whether clinical practice is aligned with best evidence and, if not, develop and test interventions to remedy this.

Translating evidence into practice is a challenging endeavour, with a time lag of 17 years commonly cited [6]. This challenge is even more problematic when the amount of evidence is growing exponentially [7]. One way to favour the evidence being disseminated to clinicians, is to involve them in the different phases of research projects, from topic selection to dissemination increasing engagement with evidence and fostering a sense of ownership.

One mechanism many professions have used to achieve this is through the creation of Practice-Based Research Networks (PBRNs). These are collaborations between clinicians and academics, aimed at fostering research in everyday clinical practice. They are a useful tool to invite clinicians in day-to-day practice to contribute to national research agenda and improvement initiatives [8,9]. They also allow clinicians to contribute data on practice-relevant topics, which in turn can identify pertinent research questions for further exploration [10]. A PBRN requires a minimum of 15 outpatient practices and/or 15 clinicians to collaborate with academic institutions to conduct research [11]. The development of a PBRN is a good way for a profession to develop a research infrastructure that goes beyond a single study and acts as a springboard for sub-studies [12]. There are several osteopathic PRBNs around the globe, including in Australia [13], New Zealand [12] and the USA [14].

Engagement in research activities among professionals can vary. There can be many reasons for this but sometimes it can be due to a lack of research training embedded in the curriculum at the time of training. The educational qualifications for osteopaths in the UK have evolved significantly over the past decades. Osteopaths completed their training with a Diploma in Osteopathy (DO), until the end of the 80s’ when the standard qualification shifted to a Bachelor of Science (BSc) degree. In the early noughties, the profession saw another transition with the introduction of a Bachelor of Osteopathy (B.Ost.) degree. This qualification remained the norm until late noughties, when many institutions began offering an integrated Master of Osteopathy (M.Ost.) [15]. These changes in degree structures were accompanied by substantial curricular modifications, particularly in the area of research methods. Consequently, a skills disparity has emerged within the profession, with practitioners’ research competencies often correlating with their graduation period. This variability in research training has implications for evidence-based practice and the profession’s overall research capacity. The development of the UK Research Network hopes to address some of these issues through minimising the research-practice translational gap, upskilling the profession in research methods, and developing the evidence base in osteopathy. This paper aims to provide data regarding the development of the NCOR Research Network, the first osteopathic Practice-Based Research Network in the UK.

## Methods

### Recruitment

The *NCOR Research Network* is the first osteopathic PBRN in the UK. A series of events were held in the year prior to its launch [16]. These events had two aims: to ensure that the *NCOR Research Network* would be meaningful to osteopaths; and to make key stakeholders and osteopaths aware of the importance of PBRNs for the professions to maximise recruitment. Four one-day events and three online webinars were delivered between October 2022 and August 2023, an exhibition stand was set up at a major international conference near London in October 2023, two articles were published in the professional association’s magazine and e-newsletters, direct contact was made with osteopathic regional groups in the UK, and a live broadcast was recorded to an audience of around 800 clinicians. Invitations were also sent by email to all osteopaths on the GOsC database who agreed to be contacted for research purposes.

### Questionnaire

A survey was used in this cross-sectional study to collect the information necessary to establish the research questions that could realistically be addressed through data collection using the PBRN. Similar surveys have been conducted with members of other PBRNs for the same purpose [12,17,18], and were used in the design of the methodology and survey for this study. Existing PBRNs were launched with a similar survey to provide data on their members. This enabled future sub-studies to target members’ preferences and settings. The questionnaire was divided into 4 sections: Section 1 contained qualifying questions to ensure that participants were eligible to take part in the survey (i.e. providing consent to take part, being registered with the General Osteopathic Council (GOsC), living and working in the UK). It also identified whether the osteopath had a clinical role and should complete Sections 2 & 3. Section 2 contained questions relating to the nature of the clinical work that the osteopath undertook (e.g., geographical location of the clinics they work in and the number of patients that they typically see per week). Section 3 contained questions about the type of patients the osteopath typically sees and how they are managed (e.g., the symptoms their patients commonly present with and what sub-groups of patients are commonly seen e.g. age groups or activities or comorbidities). Section 4 contained demographic questions about the participant (e.g. length of time in practice and professional qualifications). See Supplementary Material 1 for more details on the content and structure of the questionnaire. The data were collected via an online self-reported questionnaire, using SmartSurvey^©^ as an online platform.

### Patient and public involvement

Patients and members of the public were not involved in this phase of setting up the *NCOR Research Network*. The reason for this is that this step was to recruit osteopaths to carry out further studies. This manuscript reports on the development of this research infrastructure. Future clinical studies will involve patients and the public at an early stage of project design to ensure that projects are meaningful to them and that research is done with patients / members of the public, not on them.

### Protocol and Ethics

The protocol for this project was registered on the Open Science Framework (https://doi.org/10.17605/OSF.IO/HPWG4) before the analysis was conducted [19]. Ethical approval was received from the University College of Osteopathy Research Ethics Committee (#21122023).

### Statistical analysis

Data were imported into Microsoft Excel™. Descriptive statistics were used: dichotomous and categorical variables are presented as frequencies and percentages, and continuous variables are presented as means and standard deviations. The representativeness of the *NCOR Research Network* data was assessed by performing a chi-square test to compare its demographic characteristics to those of the broader population of GOsC-registered osteopaths.

## Results

897 participants started completing the questionnaire, 631 were fully completed and 570 participants were eligible for being included in *NCOR Research Network.* The reasons for exclusion were not being registered with the General Osteopathic Council (n=37) or participants not consenting for their data to be collected or stored (n=24).

Comparison with the GOsC registrant data revealed that the sample was not nationally representative of the UK osteopathic profession in terms of gender, age, ethnicity, and years in practice (ps < 0.005) (see supplementary material 2).

### Osteopaths’ characteristics

The median age bracket of the osteopaths was 50-59 (SD = 1.20), and 55% identified as women. They had seen patients as an osteopath for a median of 17 years (SD = 11.9). More than half of the osteopaths had a bachelor’s degree (53%), over a quarter an undergraduate master’s degree (26%), 20% a postgraduate master’s degree, and 4% a doctoral degree. The majority of their patient contact work was in private practice (71% as principals and 32% as associates), 3% worked in the NHS, and 9% were providing clinical supervision (see Table 1).

**Table 1.**
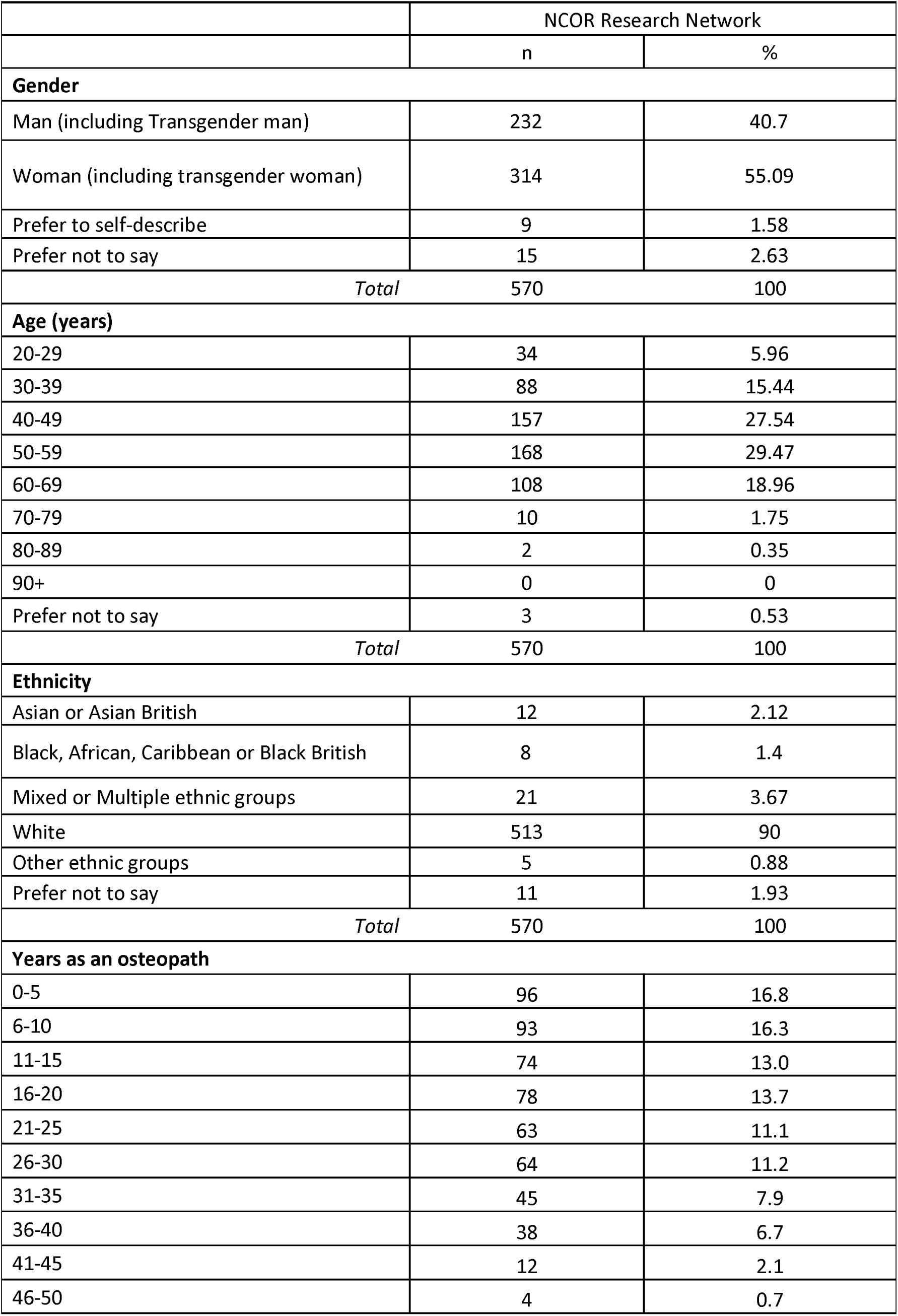
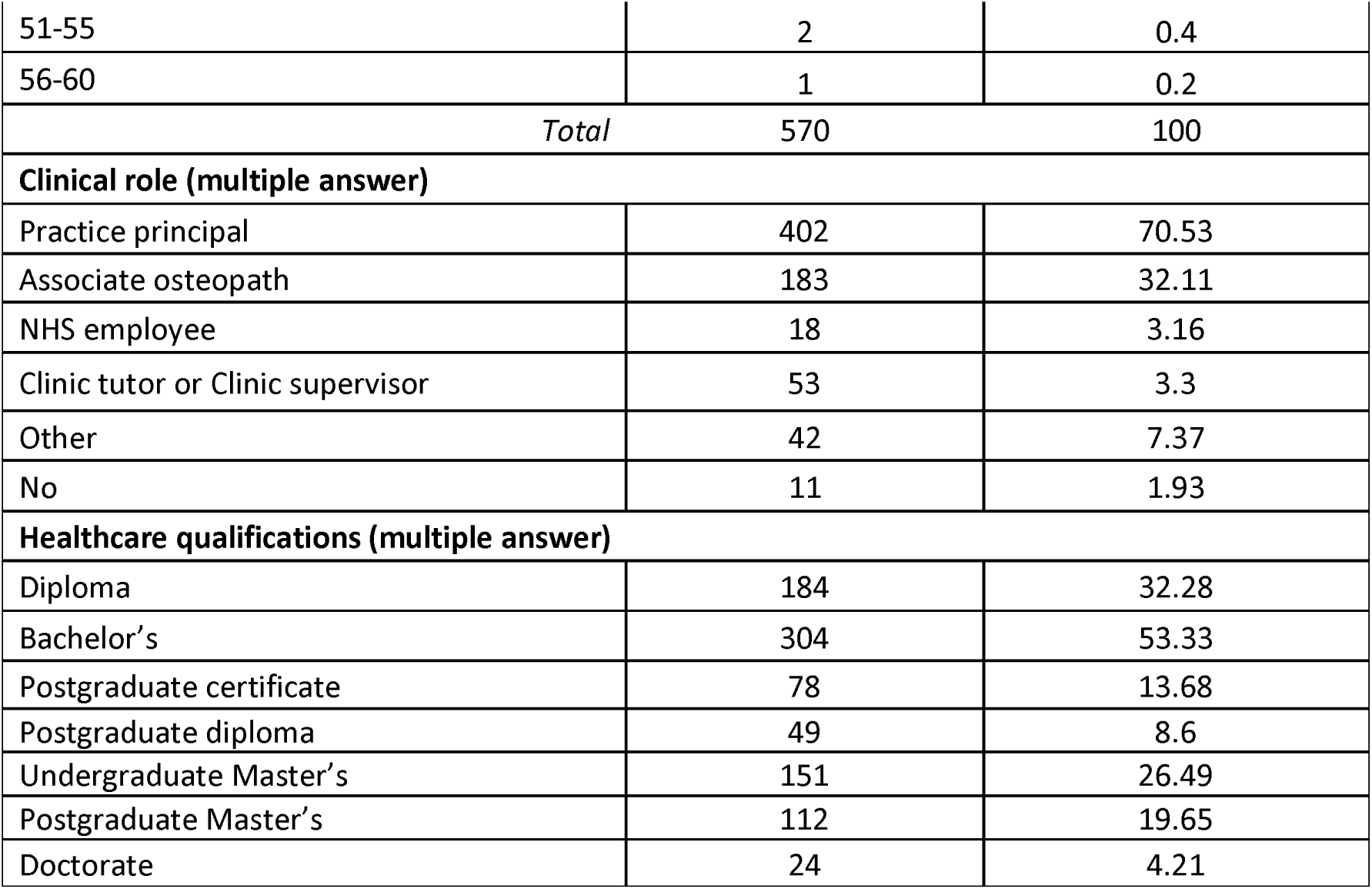
members’ demographic data.

### Osteopaths’ clinical work

Most participants were seeing patients between 20 to 29 hours (34%) or 30 to 39 hours (30%) per week, with the majority seeing 0 to 4 new patients (54%) and 32% seeing 20 to 29 follow up patients per week. Seventy-seven percent reported receiving referrals at least monthly, and the main sources were massage therapists (39%), other osteopaths (32%), GPs (29%), and health insurance companies (22%) (see Table 2).

**Table 2.** workload, sources of referral and imaging referral.

| Table 2 - workload, sources of referral and imaging referral |  |  |
| --- | --- | --- |
|  | n | % |
| Hours per week allocated for seeing patients |  |  |
| 1-9 | 27 | 4.83 |
| 10-19 | 98 | 17.53 |
| 20-29 | 192 | 34.35 |
| 30-39 | 166 | 29.7 |
| 40-49 | 60 | 10.73 |
| 50 or more | 16 | 2.86 |
| Number of new patients per week |  |  |
| 0-4 | 304 | 54.38 |
| 5-9 | 156 | 27.91 |
| 10-14 | 39 | 6.98 |
| 15-19 | 17 | 3.04 |
| 20-24 | 15 | 2.68 |
| 25 or more | 28 | 5.01 |
| Number of follow-up patients per week |  |  |
| 1-9 | 54 | 9.66 |
| 10-19 | 138 | 24.69 |
| 20-29 | 178 | 31.84 |
| 30-39 | 100 | 17.89 |
| 40-49 | 49 | 8.77 |
| 50-59 | 18 | 3.22 |
| 60 or more | 22 | 3.94 |
| Source of referrals received monthly |  |  |
| None | 129 | 23.08 |
| Acupuncturist | 112 | 20.04 |
| Chiropractor | 33 | 5.9 |
| Consultant medical doctor | 73 | 13.06 |
| Dietician | 17 | 3.04 |
| Employer or occupation health department | 34 | 6.08 |
| Health insurance company | 123 | 22 |
| Infant feeding team | 71 | 12.7 |
| GP | 162 | 28.98 |
| Massage therapist | 218 | 39 |
| Midwife | 92 | 16.46 |
| Naturopath | 16 | 2.86 |
| Nutritionist | 29 | 5.19 |
| Osteopath | 179 | 32.02 |
| Physiotherapist | 110 | 19.68 |
| Podiatrist | 60 | 10.73 |
| Psychologist or counsellor | 56 | 10.02 |
| Solicitor or other representative of the patient following an accident | 12 | 2.15 |
| Tongue tie practitioner | 77 | 13.77 |
| Other | 49 | 8.77 |
| Frequency of referral for imaging |  |  |
| Daily | 13 | 2.33 |
| Weekly | 49 | 8.77 |
| Monthly | 210 | 37.57 |
| Quarterly | 212 | 37.92 |
| Yearly | 50 | 8.94 |
| Never | 25 | 4.47 |
| Most common reason for imaging referral |  |  |
| Serious pathology identification | 211 | 39.51 |
| Musculoskeletal diagnosis confirmation | 124 | 23.22 |
| Identification of Contraindications to treatment | 30 | 5.62 |
| Patient not responding to treatment | 142 | 26.59 |
| General screening purposes | 6 | 1.12 |
| Other | 21 | 3.93 |

The majority were seeing patients in one clinic (51%), but there was a range of the number of locations, with some participants seeing patients in up to 6 or more locations (1%). Most participants were working with other healthcare professionals in their clinics (80%), the five most frequent other professionals were osteopaths (79%), massage therapists (55%), acupuncturists (37%), physiotherapists (34%), and psychologists or counsellors (32%) (see Table 3).

**Table 3.** clinical settings.

|  | n | % |
| --- | --- | --- |
| Number of clinical locations they see patients in |  |  |
| 1 | 284 | 50.81 |
| 2 | 167 | 29.87 |
| 3 | 83 | 14.85 |
| 4 | 13 | 2.33 |
| 5 | 5 | 0.89 |
| 6 or more | 7 | 1.25 |
| *Total* | 559 | 100 |
| Number of other healthcare professionals (including osteopaths) in their clinic(s) |  |  |
| 0 | 113 | 20.21 |
| 1-2 | 94 | 16.82 |
| 3-4 | 99 | 17.71 |
| 5-6 | 88 | 15.74 |
| 7-8 | 58 | 10.38 |
| 9-10 | 25 | 4.47 |
| 11 or more | 82 | 14.67 |
| *Total* | 559 | 100 |

| Type of healthcare professions in the clinic(s) |  |  |
| --- | --- | --- |
| Osteopath | 353 | 79.15 |
| Acupuncturist | 164 | 36.77 |
| Chiropractor | 57 | 12.78 |
| Consultant medical doctor | 30 | 6.73 |
| Dietician | 37 | 8.3 |
| GP | 49 | 10.99 |
| Massage therapist | 247 | 55.38 |
| Naturopath | 41 | 9.19 |
| Nutritionist | 79 | 17.71 |
| Physiotherapist | 152 | 34.08 |
| Podiatrist | 107 | 23.99 |
| Psychologist or counsellor | 141 | 31.61 |
| Other | 118 | 26.46 |

Most participants had a specialist clinical interest (56%), the top five were: cranial osteopathy (40%), chronic / persistent pain (37%), paediatrics (4 to 18 years old) (32%), sports injuries (29%), and paediatrics (under the age of 4) (27%) (see Table 4).

**Table 4.**
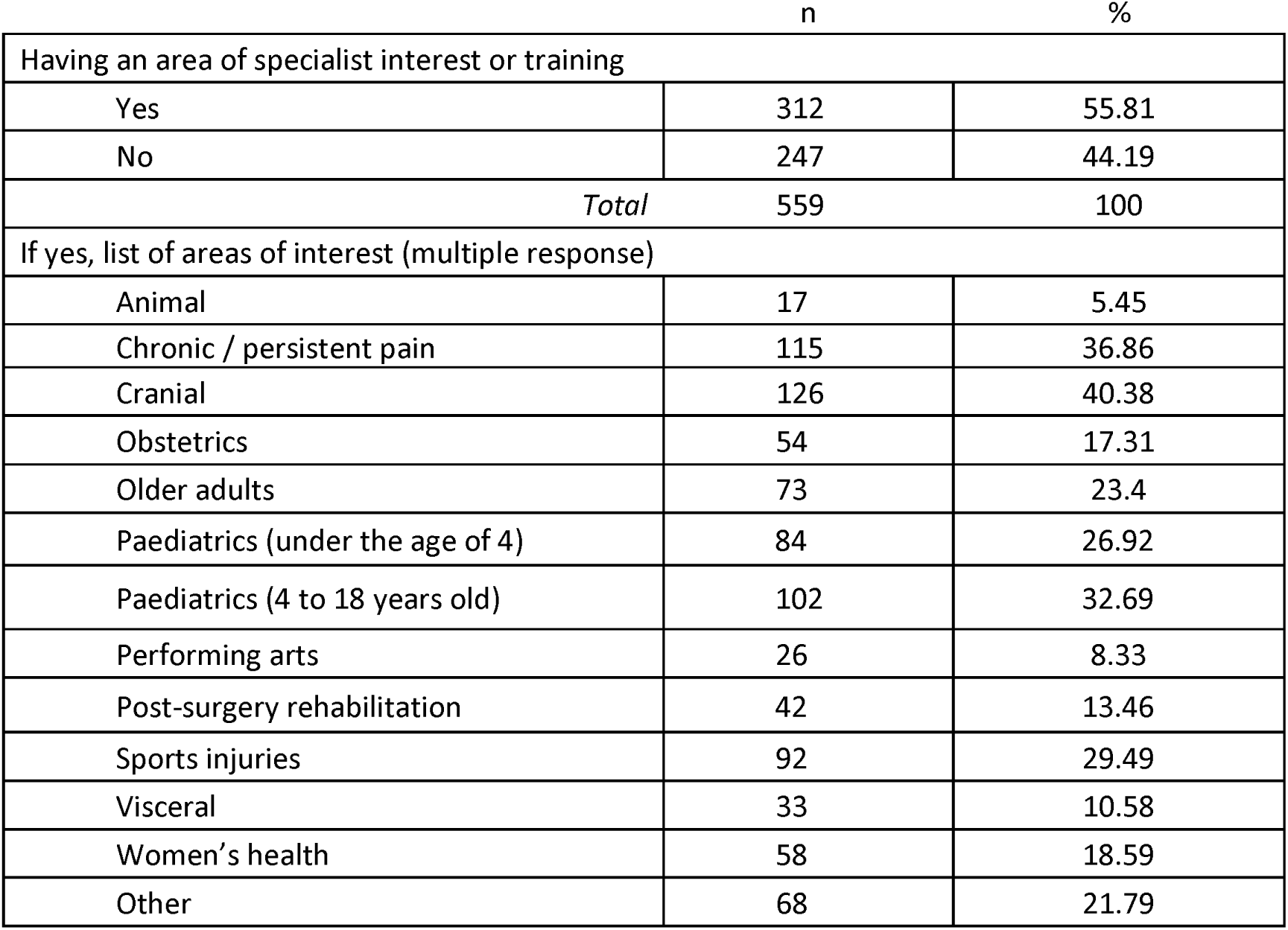
areas of special interests.

Most participants reported referring patients for diagnostic imaging quarterly (38%) or monthly (38%). The main reason was for the identification of serious pathology (40%) or when patients were not responding to treatment (27%) (see Table 2).

### Osteopaths’ patients’ characteristics

Adults 65 years of age or older were the sub-group of patients most seen by osteopaths on a regular basis (daily or weekly) (87%). People with sports-related injuries were reported as being seen regularly by 58% of osteopaths with only 2% never seeing this patient sub-group. A fifth to a third of the osteopaths saw regularly babies aged under 1 (27%), toddlers aged 1 to 3 years were seen regularly by (20%) and children aged 4 to 17 years (21%), and 50%, 48% and 9% reported never seeing these patient sub-groups respectively. A quarter of participants saw pregnant women on a regular basis, with only 5% never seeing them. Professional sports people were seen regularly by 17% of the osteopaths, with 35% reporting never seeing this subgroup of patients (see Table 5).

**Table 5.** patient groups.

| Table 5 - patient groups |  |  |  |  |  |  |
| --- | --- | --- | --- | --- | --- | --- |
|  | Daily<br>%<br>n | Weekly<br>%<br>n | Monthly<br>%<br>n | Quarterly<br>%<br>n | Yearly<br>%<br>n | Never<br>%<br>n |
| Adults 65 years of age or older | 45.62%<br>255 | 41.32%<br>231 | 9.66%<br>54 | 2.50%<br>14 | 0.54%<br>3 | 0.36%<br>2 |
| Pregnant women (regardless of their complaint) | 3.22%<br>18 | 21.47%<br>120 | 31.66%<br>177 | 26.30%<br>147 | 12.34%<br>69 | 5.01%<br>28 |
| People with sports-related injuries | 15.03%<br>84 | 42.93%<br>240 | 28.80%<br>161 | 9.48%<br>53 | 1.61%<br>9 | 2.15%<br>12 |
| Professional sports people | 2.86%<br>16 | 14.13%<br>79 | 15.21%<br>85 | 15.21%<br>85 | 17.89%<br>100 | 34.70%<br>194 |
| Babies aged under 1 | 7.87%<br>44 | 19.32%<br>108 | 10.02%<br>56 | 6.08%<br>34 | 6.26%<br>35 | 50.45%<br>282 |
| Toddlers aged 1 to 3 years | 3.40%<br>19 | 16.10%<br>90 | 11.45%<br>64 | 9.30%<br>52 | 11.63%<br>65 | 48.12%<br>269 |
| Children aged 4 to 17 years | 2.33%<br>13 | 18.78%<br>105 | 28.44%<br>159 | 30.59%<br>171 | 10.91%<br>61 | 8.94%<br>50 |
| People with road traffic accident injuries | 2.86%<br>16 | 17.35%<br>97 | 23.97%<br>134 | 32.02%<br>179 | 17.89%<br>100 | 5.90%<br>33 |
| People requiring post-surgical rehabilitation | 3.76%<br>21 | 17.53%<br>98 | 23.08%<br>129 | 24.51%<br>137 | 19.32%<br>108 | 11.81%<br>66 |
| Non-English-speaking people | 2.68%<br>15 | 15.03%<br>84 | 10.91%<br>61 | 20.75%<br>116 | 25.40%<br>142 | 25.22%<br>141 |

### Patients’ symptoms

Participants reported mostly seeing patients with musculoskeletal complaints: 71% reported that 75% to 100% of their patients consulted with MSK symptoms as their main complaint (see Table 6). Participants reported the frequency they were seeing patients (including new and follow-up) for different complaints. The only complaint that was most frequently seen on a daily basis was low back pain with or without radiculopathy (56%). Complaints that were mostly seen weekly were knee pain (57%), hip pain (57%) shoulder pain (54%), headaches (51%), mid or upper back pain (50%), and neck pain with or without radiculopathy (49%). Complaints that were mostly seen monthly were elbow pain (40%) and foot pain (35%). The complaint that was mostly seen quarterly was hand pain (30%). Two complaints were mostly never seen: non-musculoskeletal paediatric complaints (48%) and non-musculoskeletal adult complaints (31%) (see Table 7).

**Table 6.**
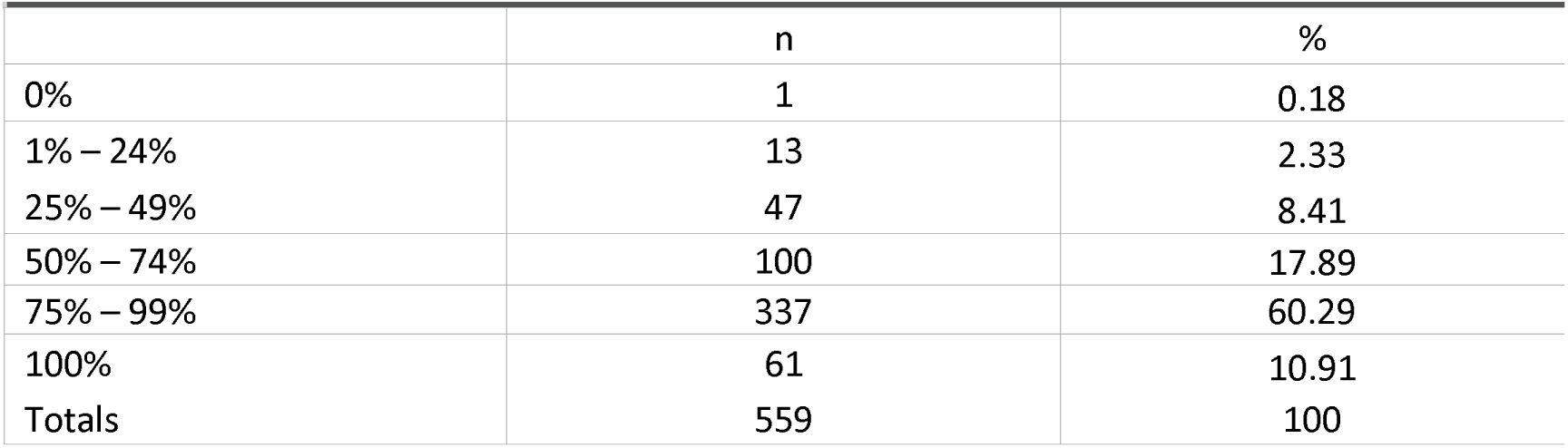
percentage of patients with musculoskeletal pain as their main complaint.

|  | n | % |
| --- | --- | --- |
| 0% | 1 | 0.18 |
| 1% – 24% | 13 | 2.33 |
| 25% – 49% | 47 | 8.41 |
| 50% – 74% | 100 | 17.89 |
| 75% – 99% | 337 | 60.29 |
| 100% | 61 | 10.91 |
| Totals | 559 | 100 |

**Table 7.**
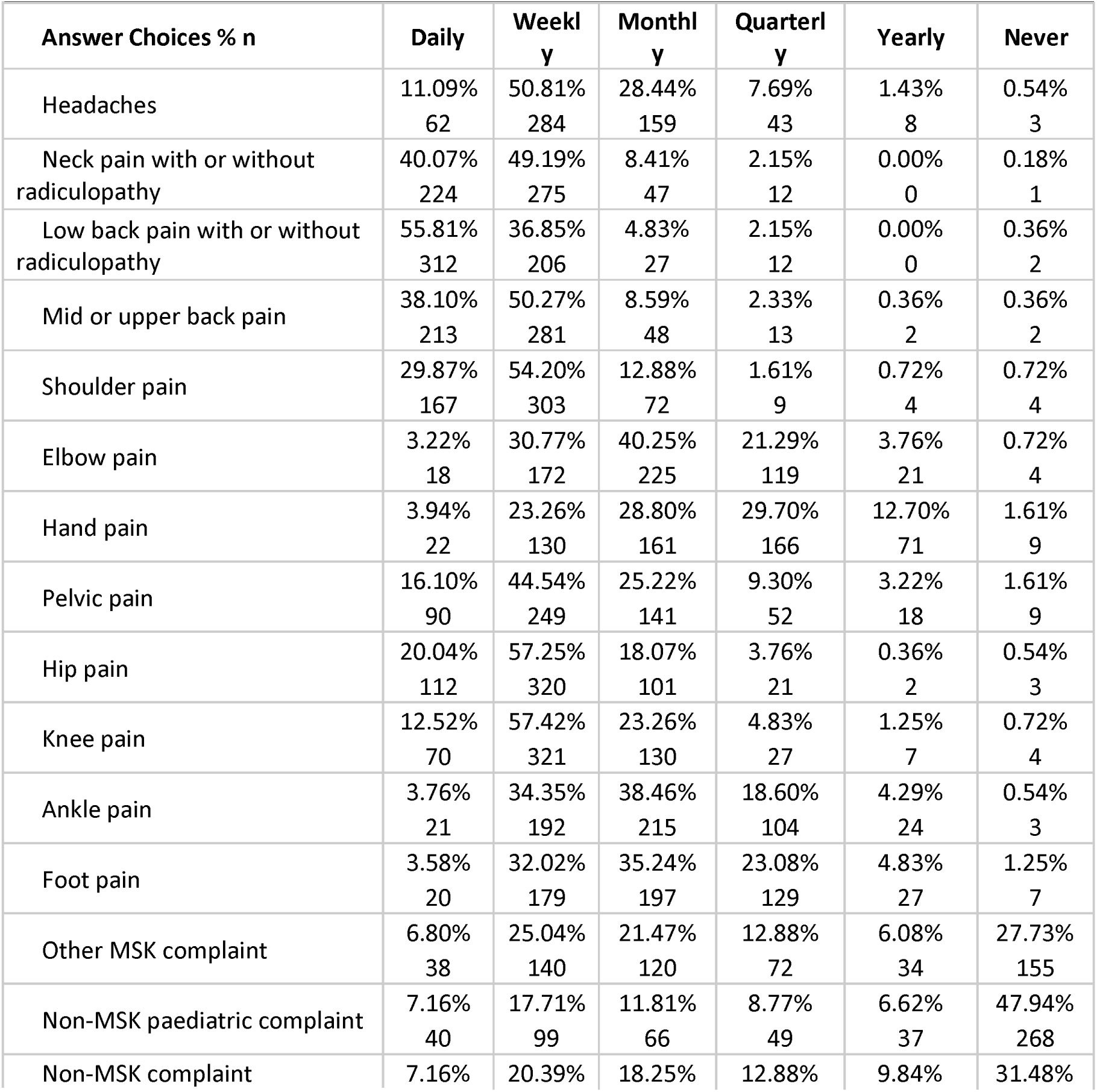
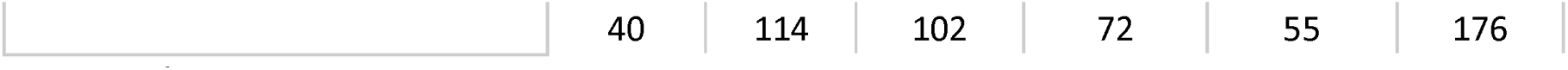
complaints frequency.

### Osteopathic management

Participants reported the frequency they were using different techniques or approaches in patient management. The approaches that were mostly used daily were: Spinal articulation or mobilisation (79%), Soft tissue massage (78%), Exercise recommendation (74%), Muscle Energy Technique (MET) or Proprioceptive Neuromuscular Facilitation (PNF) (57%), High Velocity Thrust (HVT) or spinal manipulation/adjustment (50%), General Osteopathic Treatment (GOT) or General Body Adjustment (GBA) (33%) and cranial osteopathy (32%). Several approaches were mostly reported as never being used by the majority of the participants: Intervertebral Differential Dynamics (IDD) therapy or Intermittent Sustained Spinal Traction (ISST) (89%), Transcutaneous Electrical Nerve Stimulation (TENS) or Electrical Muscle Stimulation (EMS) (84%), Extracorporeal Shockwave Therapy (ESWT) (83%), Laser therapy (85%), Instrument assisted soft-tissue (78%), Ultrasound (77%), and Dry needling or acupuncture (54%). Seventy four percent reported never using any other approaches than the ones listed. Some approaches were variable in the frequency they were used: Strain-Counterstrain or Facilitated Positional Release were used weekly by 26% of the participants whilst 25% never used them; neurodynamics or flossing were used weekly by 28% of participants whilst 33% never used them (see Table 8).

**Table 8.**
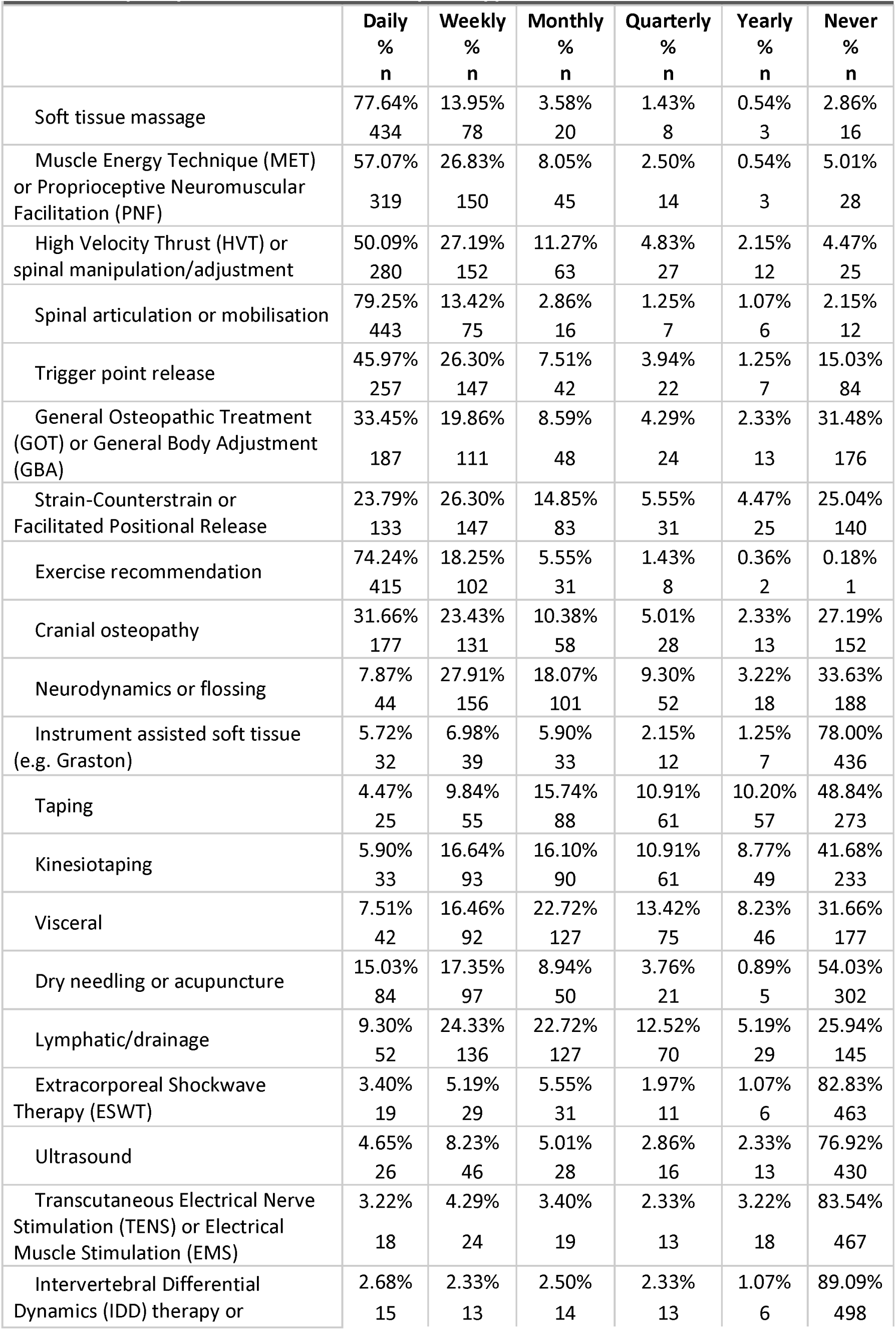
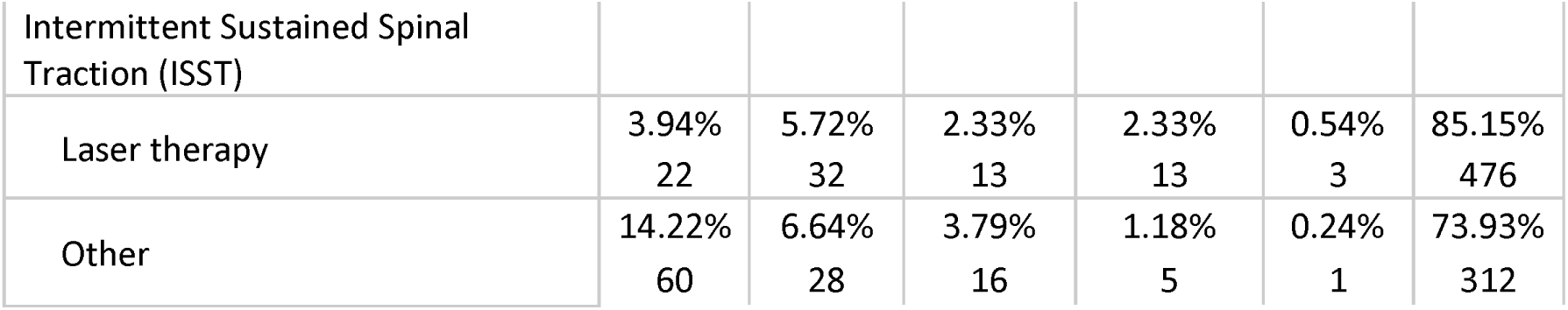
frequency of use of different techniques / approaches.

In terms of other strategies to support patient self-management, participants reported that they mostly discussed these approaches on a daily basis: general physical activity (not specific MSK exercise rehabilitation) (65%), stress management (45%), medication (including for pain/inflammation) (43%), occupational health and safety or ergonomics (42%), and diet or nutrition (36%). On a weekly basis: nutritional supplements (including vitamins, minerals, herbs) (30%), and smoking, drugs or alcohol cessation (27%). Infant feeding advice was never discussed by 50% of the participants, and other health promotion advice or education was never discussed by 37% of the participants (see Table 9).

**Table 9.** frequency of other management strategies.

| Table 9 - frequency of other management strategies |  |  |  |  |  |  |
| --- | --- | --- | --- | --- | --- | --- |
| Answer Choices | Daily<br>%<br>n | Weekly<br>%<br>n | Monthly<br>%<br>n | Quarterly<br>%<br>n | Yearly<br>%<br>n | Never<br>%<br>n |
| Diet or nutrition | 35.96%<br>201 | 35.06%<br>196 | 18.96%<br>106 | 6.62%<br>37 | 1.61%<br>9 | 1.79%<br>10 |
| Smoking, drugs or alcohol | 21.65%<br>121 | 27.19%<br>152 | 23.26%<br>130 | 13.42%<br>75 | 7.51%<br>42 | 6.98%<br>39 |
| General physical activity (not specific MSK exercise rehab) | 65.12%<br>364 | 24.87%<br>139 | 8.41%<br>47 | 1.25%<br>7 | 0.36%<br>2 | 0.00%<br>0 |
| Occupational health and safety or ergonomics | 42.40%<br>237 | 39.71%<br>222 | 11.27%<br>63 | 4.29%<br>24 | 1.07%<br>6 | 1.25%<br>7 |
| Stress management | 44.72%<br>250 | 38.10%<br>213 | 12.70%<br>71 | 3.40%<br>19 | 0.54%<br>3 | 0.54%<br>3 |
| Nutritional supplements (including vitamins, minerals, herbs) | 18.96%<br>106 | 30.05%<br>168 | 23.97%<br>134 | 11.99%<br>67 | 4.65%<br>26 | 10.38%<br>58 |
| Medication (including for pain/inflammation) | 43.47%<br>243 | 36.14%<br>202 | 12.52%<br>70 | 3.22%<br>18 | 1.61%<br>9 | 3.04%<br>17 |
| Infant feeding advice | 9.30%<br>52 | 15.74%<br>88 | 10.38%<br>58 | 7.51%<br>42 | 7.51%<br>42 | 49.55%<br>277 |
| Pain science education | 33.09%<br>185 | 29.70%<br>166 | 18.25%<br>102 | 6.62%<br>37 | 3.58%<br>20 | 8.77%<br>49 |
| Other health promotion advice or education (please specify below) | 25.11%<br>114 | 23.13%<br>105 | 8.81%<br>40 | 4.85%<br>22 | 0.66%<br>3 | 37.44%<br>170 |

The majority of the participants had positive views regarding evidence-based practice. Participants reported lacking research training, experience or skills beyond their undergraduate training; and lacking spare time for research related to osteopathic practice (see Table 10).

**Table 10.**
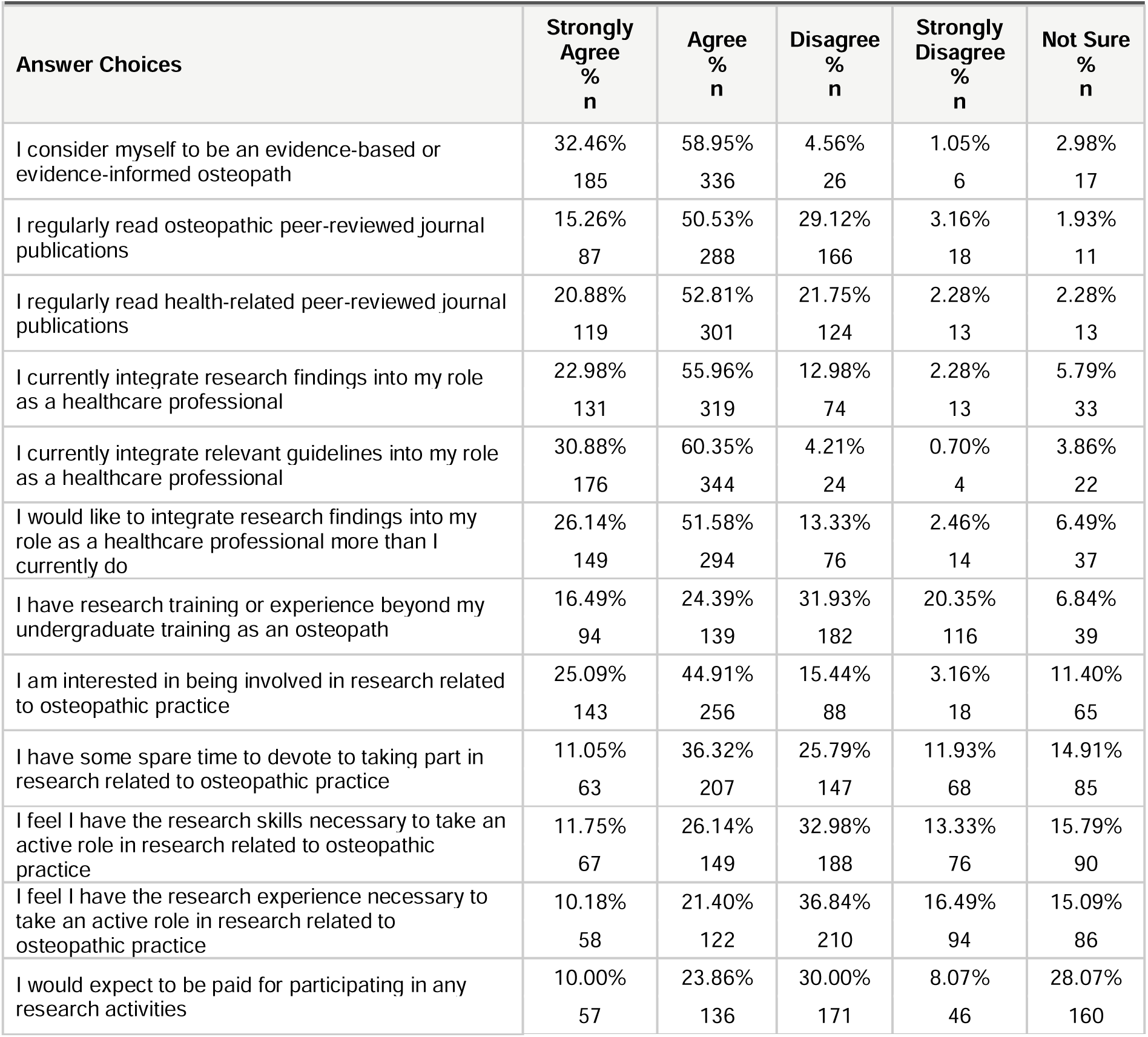
Members’ views on Evidence-Based Practice.

## Discussion

The aim of this paper was to provide data regarding the development of the *NCOR Research Network*, the first osteopathic Practice-Based Research Network in the UK. Several interesting key findings were identified.

Clinical practice reported by *NCOR Research Network* osteopaths demonstrates substantial alignment with current clinical guidelines. For instance, the criteria reported by osteopaths for referring patients for diagnostic imaging are consistent with international guidelines for musculoskeletal care [20]. Moreover, the incorporation of self-management strategies in treating musculoskeletal complaints reflects adherence to national guidance [21,22]. These practices suggest a growing trend towards evidence-informed care within the osteopathic profession. This shift is further corroborated by longitudinal data on practitioners’ attitudes. Between 2014 and 2020, there was an increase in the proportion of osteopaths who agreed or strongly agreed that evidence-based practice improves patient care, rising from 38% to 50% [23,24]. This trend indicates a gradual but significant change in the profession’s perspective on the value of evidence-based approaches. Future research endeavours could productively focus on exploring the impact of the *NCOR Research Network* on osteopaths’ sense of engagement with research and the ongoing evolution of attitudes towards evidence-informed practice. This could provide insights into the factors driving the profession’s increasing embrace of evidence-based methodologies and identify potential barriers or facilitators to this transition.

This is the first report including UK osteopaths’ qualifications in a research study. More than half of the osteopaths had a bachelor’s degree, nearly half had a master’s level degree and 4% a doctoral degree related to healthcare. This is a higher number than for osteopaths practising in Germany, Italy, Switzerland, and Belgium-Netherlands-Luxembourg, but lower than in Australia [25]. The percentage of osteopaths with a doctoral degree is higher than in other osteopathic PBRNs (e.g. 0.5% in ORION [13]). Despite the overall high academic qualifications of our osteopaths, the main challenges they reported facing regarding taking part in research were a lack of research skills, and a lack of time. This identifies a need to develop research training for *NCOR Research Network* osteopaths to facilitate their engagement with research and ensure a positive research culture.

The majority of *NCOR Research Network* work with other healthcare professionals. This is different from a previous survey of osteopaths in the UK that found that 64% were working alone [1], compared to 20% in our participants. This difference may be due to our sample not representing the profession or related to changes that may have happened since the above-mentioned survey data was collected in 2018. The self-selecting nature of the *NCOR Research Network* survey participants may have attracted those who are more inclined to want to work with others, given the ‘network’ focus of the survey.

*NCOR Research Network* osteopaths reported using a variety of manual therapy approaches that have previously been identified as commonly used by osteopaths [25]. They reported also discussing a range of health-related topics with their patients, including physical activity, stress management, medication, occupational health and safety or ergonomics, diet or nutrition, smoking, drugs or alcohol. These types of discussions are consistent with national initiatives for healthcare professionals such as Making Every Contact Count [26], however, there is a need to assess the nature of these discussions and possibly to assess the usefulness of existing interventions to support clinicians treating MSK conditions.

Despite the lack of representativeness of the *NCOR Research Network*sample, the clinical data reported was consistent with data collected in the UK in 2019 [1]. The similar characteristics included the predominant subgroup of patients being 65 years or older, musculoskeletal complaints as the primary reason for consultation, and consistency in techniques and approaches implemented in patient management. The *NCOR Research Network*osteopaths reported more frequent use of general physical activity advice, stress management techniques, medication discussions, and diet and nutrition discussions compared to osteopaths in 2019. These differences may be attributed to either the lack of representativeness of the *NCOR Research Network*sample or potential shits in osteopathic practice over the past five years. Several aspects of osteopathic practice remain underexplored, highlighting a pressing need for additional data collection and analysis: including the decision-making process that leads patients to seek osteopathic care, or the factors influencing the discontinuation of osteopathic care. A well-designed system will be required to prioritise efficiency in data collection, as osteopaths have expressed concerns about time-consuming nature of participation in research. Striking a balance between comprehensive data gathering and minimising the burden on osteopaths will be crucial for future studies in this field.

### Limitations

The sample in our study is not representative of the profession in the UK when compared to the GOsC registrant data. The recruitment strategy used different methods, including events in different parts of the UK, to promote the *NCOR Research Network* to as many of the profession as possible. Whilst the response rate was above the requirements for the setup of a PBRN, we may have to consider promoting *NCOR Research Network* to specific groups to help developing representativeness, particularly those who have been in practice for less than 10 years. Another limitation is that the survey data were self-reported by osteopaths, so the findings may be impacted by recall or social desirability biases.

### Strengths

The survey was widely promoted to the osteopathic profession through various channels available in the UK. These included: the regulatory body (GOsC), the professional organisation (Institute of Osteopathy), professional publications, and face-to-face and online presentations to diverse groups. Prior to initiating recruitment, qualitative work was conducted to explore key issues in depth. This preliminary research helped identify pertinent topics for inclusion in the survey’s development.

The survey development was informed by similar surveys that had been used for other osteopathic PBRN set ups [12], and by other osteopathic datasets in the UK, including PROMs [27] and the Standardised Data Collection tool [28], to allow comparability of data.

## Conclusion

*NCOR Research Network* provides a useful infrastructure to support the development of research in osteopathy and related fields through collaboration with other healthcare professionals and researchers. It is also an innovative approach in the UK to foster collaboration between osteopaths and academics. It will help to better understand what is done in real-world practice [29], by collecting data from clinicians (e.g. about their patients, their management strategies, or the number of sessions), from patients (e.g. using Patient Reported Outcome Measures or Patient Reported Experience Measures), or combining both patients’ and clinicians’ data [27,30]. A range of study designs can be employed, including observational studies, pragmatic clinical studies, and qualitative research [12,31]. Osteopaths in the UK who would like to join NCOR Research Network can find further information on https://ncor.org.uk/PBRN/.

## Data Availability

Data sharing statement: data are presented in the tables of the manuscript. For any queries, please contact NCOR directly.

## Data sharing statement

data are presented in the tables of the manuscript. For any queries, please contact NCOR directly.

## Author contributions

JDR, CF, and DB conceptualised and designed the study. DB registered the protocol. DB led the data collection. DB and JDR conducted the data analysis. JDR drafted the first version of the manuscript. All authors (JDR, CF, and DB) critically revised the manuscript for important intellectual content, provided substantial input to subsequent drafts, and approved the final version. All authors agree to be accountable for all aspects of the work in ensuring that questions related to the accuracy or integrity of any part of the work are appropriately investigated and resolved.

## Acknowledgements section

We would like to thank Prof Alice Kongsted (University of Southern Denmark) and Dr Amie Steel (University of Technology Sydney) for their generous support by sharing their experience and expertise with setting up Practice-Based Research Networks.

