## Supplemental file 1 for "Development of a national osteopathic practice-based research network: the NCOR Research Network"

### NCOR Research Network Survey

#### 1. Participant Information

**Please read the following information before starting the survey, if you have any questions or require further information, please email Daniel Bailey, at. By continuing to the survey, you are providing consent for us to collect and store your information as described below.** 

**1.Study title**
NCOR Research Network Members’ Survey
**2. Invitation**
We would like to invite you to join the NCOR Research Network, the UK’s first Practice Based Research Network (PBRN) for osteopathy. PBRNs offer unique opportunities for clinicians in day-to-day practice to contribute to research by providing data on practice-relevant topics. To identify feasible research topics, we first need to establish the demographics of the Network’s members. We are therefore asking you to complete a survey collecting information about your role as an osteopath and if relevant, information about your clinical practice. Please feel free to talk to others about the study if you wish and contact us if anything is unclear.
**3. What is the purpose of the study?**
The aim of this study is to establish the research capacity of the NCOR Research Network, specifically, the clinical environments, patient demographics, and clinician demographics of its members.
**4. Why have I been invited?**
You have been invited because you are a registered osteopath in the UK. As the Network intends to inform osteopathic practice in the UK, its members must be UK-registered osteopaths. Consequently, to be eligible to complete this survey, you must be a UK-based osteopath registered with the General Osteopathic Council (GOsC). You do not need to have a clinical role seeing patients to be eligible to complete the survey or join the Network.
**5.   Do I have to take part?**
No, your involvement in this study is purely voluntary and of course you do not have to take part if you do not want to. You will have three months from receipt of the invitation to complete the survey and be included in the prize draw.  After this time, you can still join the Network by submitting your completed survey, but you will not be included in the prize draw and your data will not be included in the initial analysis, although it will still be stored for use with the Network.  You can stop the survey at any time and contact the research team if you have any questions or concerns. If you decide to withdraw from the study during the three-month period following the invitation, you can choose to have any information you have submitted removed from the study and destroyed. However, if you decide to withdraw from the study after this time and have already submitted the survey, you will not be able to withdraw your information. Once you have submitted the survey, there is no obligation to take part in any further activity, although you will be part of the NCOR Research Network and invited to take part in future research activities.  You are free to leave the Network at any time.
**6. What will happen to me if I take part?**
You will be invited to fill in an online survey about you and your professional environment. The questionnaire will take approximately 15-20 minutes to complete, although you do not have to complete it all in one go.  You may then be invited by NCOR to take part in future studies, but completing this survey does not commit you to any future projects.
**7. What do I have to do?**
If you would like to take part in the study, please read the rest of this page and then click the 'Next Page' button at the bottom of the page.
**8. What are the possible benefits of taking part?**
If you decide to take part in this project, you will benefit from the opportunities available to Research Network members, including involvement in setting of research priorities and access to free continuing professional development including in research skills.  All surveys completed and returned within three months will be entered into a prize draw, with the winner receiving a £100 Amazon voucher. 
**9. What are the possible disadvantages and risks of taking part?**
There are no known risks to you from taking part. The main disadvantage is the time required to participate, but no financial disadvantages are expected.
**10. What if there is a problem?**
You should contact the lead researcher, Daniel Bailey at. If you still have a problem, you should contact NCOR’s Director, Dr Jerry Draper-Rodi at.
**11. Will my taking part in the study remain confidential?**
All the information you give us will be treated in the strictest confidence and used only for the purpose of the Research Network. Completed surveys will be anonymised before being analysed. All possible steps to ensure confidentiality will be taken and any data entering the public domain (e.g. journal publications) will be reviewed to ensure identification of individual participants is not possible. The data will only be accessible to the NCOR research team.
**12. What will happen to the results from the study?**
The results of the survey will eventually be disseminated through journal publications and presentation at conferences. This will include a video presentation to Network members.
**13. Who is organising the research?**
The research is organised by the National Council of Osteopathic Research, and the study was approved by the University College of Osteopathy (UCO) Research Ethics Committee.

**Thank you for taking the time to read the information sheet, if you would like to download a copy, please click the** [**link**](https://ncor.org.uk/pbrn/)**.  By continuing to the survey, you are providing consent for us to collect and store your information as described above.** 

#### 2. Consent

##### 1.  Do  you consent for us to collect and store your information as described on the previous page?

|  | Yes, I consent for you to collect and store my information |
| --- | --- |
|  | No, I do not consent for you to collect and store my information |

#### 3. Qualifying Question

##### 2. Are you registered with the General Osteopathic Council (GOsC) and living and working in the UK?

|  | Yes |
| --- | --- |
|  | No |

#### 4. About your clinical work

##### 3. Do you have a clinical role seeing patients as an osteopath? (Please select all that apply)

No

Practice principal

Associate osteopath

NHS employee

Clinic tutor or clinical supervisor

Other (please specify):

#### 5. About your clinical work

##### 4. What is your role related to osteopathy? (Please select all that apply)

Practice management/administration

Teaching in an osteopathic educational institution

Teaching in another healthcare setting

Education management

Providing post-graduate training

Research in an osteopathic educational institution

Research in another osteopathic setting

Research in a non-osteopathic setting

Healthcare governance or regulation

Healthcare management in the NHS

Committee member, e.g., GOsC fitness to practice

Mentoring

Other (please specify):

#### 6. About your clinical work

##### 5. Do you have any healthcare roles other than seeing patients? (Please select all that apply)

No

Practice management/administration

Teaching in an osteopathic educational institution

Teaching in another healthcare setting

Education management

Providing post-graduate training

Research in an osteopathic educational institution

Research in another osteopathic setting

Research in a non-osteopathic setting

Healthcare governance or regulation

Healthcare management in the NHS

Committee member, e.g., GOsC fitness to practice

Mentoring

Other (please specify):

#### 7. About your clinical work

##### 6. How many different clinical locations do you see patients in? (Please select one option only)

|  | 1 |
| --- | --- |
|  | 2 |
|  | 3 |
|  | 4 |
|  | 5 |
|  | 6 or more |

#### 8. About your clinical work

##### 7. In which area(s) of the UK do you see patients in? (Please select all that apply)

|  | Northeast England |
| --- | --- |
|  | Northwest England |
|  | Yorkshire and the Humber |
|  | East Midlands |
|  | West Midlands |
|  | East of England |
|  | London |
|  | Southeast England |
|  | Southwest England |
|  | Eastern Scotland |
|  | Highlands and Islands |
|  | Northeastern Scotland |
|  | West Central Scotland |
|  | Southern Scotland |
|  | West Wales and The Valleys |
|  | East Wales |
|  | Northern Ireland |

#### 9. About your clinical work

##### 8. Which digital patient record keeping system(s) do you use in the clinic(s) you see patients in? (Please select all that apply)

None, I use paper records

Cliniko

Halaxy

Jane

Power Diary

Private Practice Software

TM2 or TM3

NHS digital records system

Other (please specify):

#### 10. About your clinical work

##### 9. Which of the following settings do you see patients in? (Please select all that apply)

Private practice as the only clinician

Private practice with other osteopaths only

Private practice with other osteopaths and other healthcare professionals

Private practice as the only osteopath but with other healthcare professionals

Home visits

Virtually using video calls or telephone calls

GP practice seeing privately funded patients

GP practice seeing NHS funded patients

NHS hospital

Private hospital

NHS community provider

Other (please specify):

#### 11. About your clinical work

##### 10. How many other healthcare professionals (including osteopaths) work in the clinic(s) you see patients in (including those that work at different times to you)? (Please select one option only)

|  | 0 |
| --- | --- |
|  | 1– 2 |
|  | 3 – 4 |
|  | 5 – 6 |
|  | 7 – 8 |
|  | 9 – 10 |
|  | 11 or more |

#### 12. About your clinical work

##### 11. What type of health care professionals work in the clinic(s) you see patients in (including those that work at different times to you)? (Please select all that apply)

Osteopath

Acupuncturist

Chiropractor

Consultant Medical Doctor (e.g., Rheumatologist)

Dietician

General Practitioner (GPs)

Massage therapist

Naturopath

Nutritionist

Physiotherapist

Podiatrist

Psychologist or counsellor

Other (please specify):

#### 13. About your clinical work

##### 12. How many hours per *****week***** do you normally allocate for seeing patients, regardless of how many appointments are actually booked? (Please select one option only)

|  | 1 – 9 |
| --- | --- |
|  | 10 – 19 |
|  | 20 – 29 |
|  | 30 – 39 |
|  | 40 – 49 |
|  | 50 or more |

#### 14. About your clinical work

##### 13. Based on the last *****three months*****, how many *****new***** patients (patients that have never been to the clinic you are working in before) do you estimate that you see per week across all the clinics you work at? (Please select one option only)

|  | 0 – 4 |
| --- | --- |
|  | 5 – 9 |
|  | 10 – 14 |
|  | 15 – 19 |
|  | 20 – 24 |
|  | 25 or more |

#### 15. About your clinical work

##### 14. Based on the last *****three***** *****months*****, how many follow-up patients (patients that have been to the clinic you are working in before) do you estimate that you see per *****week***** across all the clinics you work at? (Please select one option only)

|  | 1 – 9 |
| --- | --- |
|  | 10 – 19 |
|  | 20 – 29 |
|  | 30 – 39 |
|  | 40 – 49 |
|  | 50 – 59 |
|  | 60 or more |

#### 16. About your clinical work

##### 15. Would you describe yourself as having a special area of interest or training that means you typically see more of a specific type of patient or use a particular approach/technique?

|  | Yes |
| --- | --- |
|  | No |

#### 17. About your clinical work

##### 16. How would you describe your area of special interest? (Please select all that apply)

Animal

Chronic/persistent pain

Cranial

Obstetrics

Older adults (over the age of 65)

Paediatrics (children 4 to 18 years of age)

Paediatrics (children under 4 years of age)

Performing arts

Post-surgical rehabilitation

Sports injuries

Visceral

Women’s health

Other (please specify):

#### 18. About your clinical work

##### 17. What additional training do you have in the areas of specialist interest you selected in the previous question? (Please select all that apply)

None

Shadowing of experienced colleague working with this patient group/field

Personal research or wider reading regarding this patient group/field

Personal experience in clinic with this patient group/field

Completion of training courses related to this patient group/field

Completion of specific training courses for this patient group/field

Bachelor’s degree (E.g., BSc) related to this patient group/field

Post-graduate Certificate (PgCert) related to this patient group/field

Post-graduate Diploma (PgDip) related to this patient group/field

Post-graduate Masters (MSc or MA) related to this patient group/field

Master of Research (MRes) related to this patient group/field

Master of Philosophy (MPhil) related to this patient group/field

Professional Doctorate (ProfDoc) related to this patient group/field

Philosophy Doctorate (PhD) related to this patient group/field

Other (please specify):

#### 19. About your patients

##### 18. Which languages are commonly spoken by the patients that you see (including all the clinics you work in)? (Please select all that apply)

English

Welsh

Gaelic and Scots

Irish and Ulster Scots

Cornish

Punjabi

Urdu

Polish

Other (please specify):

#### 20. About your patients

##### 19. What percentage of patients that you are currently seeing for treatment, do you estimate presented with musculoskeletal pain as their main complaint? (Please select one option only)

|  | 0% |
| --- | --- |
|  | 1% – 24% |
|  | 25% – 49% |
|  | 50% – 74% |
|  | 75% – 99% |
|  | 100% |

#### 21. About your patients

##### 20. Over the course of the *****last year*****, how frequently do you estimate that you have referred a patient to another healthcare professional, including their General Practitioner (GP)? (Please select one option only)

|  | Daily |
| --- | --- |
|  | Weekly |
|  | Monthly |
|  | Quarterly |
|  | Yearly |
|  | Never |

#### 22. About your patients

##### 21. Do you regularly (at least on a monthly basis) have patients referred to you from other health care professionals? (Please select all that apply)

No

Acupuncturist

Chiropractor

Consultant Medical Doctor (e.g., Rheumatologist)

Dietician

Employer or occupation health department

Health insurance company

Infant Feeding Team

General Practitioner (GP)

Massage therapist

Midwife

Naturopath

Nutritionist

Osteopath

Physiotherapist

Podiatrist

Psychologist or counsellor

Solicitor or other representative of the patient following an accident

Tongue Tie Practitioner

Other (please specify):

#### 23. About your patients

##### 22. Over the course of the *****last year*****, how frequently do you estimate that you have referred a patient for diagnostic imaging? (Please select one option only)

|  | Daily |
| --- | --- |
|  | Weekly |
|  | Monthly |
|  | Quarterly |
|  | Yearly |
|  | Never |

#### 24. About your patients

##### 23. What is the most common reason that you refer patients for diagnostic imaging? (Please select one option only)

To identify a serious underlying pathology (e.g., cancer, infection, trauma, or inflammatory disease)

To confirm an MSK diagnosis

To identify possible contraindications to treatment

The patient is not responding to treatment

For general screening purposes

Other (please specify):

#### 25. About your patients

##### 24. Please indicate the frequency that you typically see patients (including new and follow-up) with the following complaints. (Please select one response for each complaint)

|  | Daily | Weekly | Monthly | Quarterly | Yearly | Never |
| --- | --- | --- | --- | --- | --- | --- |
| Headaches |  |  |  |  |  |  |
| Neck pain with or without radiculopathy |  |  |  |  |  |  |
| Low back pain with or without radiculopathy |  |  |  |  |  |  |
| Mid or upper back pain |  |  |  |  |  |  |
| Shoulder pain |  |  |  |  |  |  |
| Elbow pain |  |  |  |  |  |  |
| Hand pain |  |  |  |  |  |  |
| Pelvic pain |  |  |  |  |  |  |
| Hip pain |  |  |  |  |  |  |
| Knee pain |  |  |  |  |  |  |
| Ankle pain |  |  |  |  |  |  |
| Foot pain |  |  |  |  |  |  |
| Other MSK complaint, please specify below |  |  |  |  |  |  |
| Non-MSK paediatric complaint, please specify below |  |  |  |  |  |  |
| Non-MSK complaint, please specify below |  |  |  |  |  |  |

Comment:

#### 26. About your patients

##### 25. Please indicate the frequency that you treat patients (including new and follow-up) from the following subgroups.  (Please select one response for each subgroup)

|  | Daily | Weekly | Monthly | Quarterly | Yearly | Never |
| --- | --- | --- | --- | --- | --- | --- |
| Adults 65 years of age or older |  |  |  |  |  |  |
| Pregnant women (regardless of their complaint) |  |  |  |  |  |  |
| People with sports-related injuries |  |  |  |  |  |  |
| Professional sports people |  |  |  |  |  |  |
| Babies aged under 1 |  |  |  |  |  |  |
| Toddlers aged 1 to 3 years |  |  |  |  |  |  |
| Children aged 4 to 17 years |  |  |  |  |  |  |
| People with road traffic accident injuries |  |  |  |  |  |  |
| People requiring post-surgical rehabilitation |  |  |  |  |  |  |
| Non-English-speaking people |  |  |  |  |  |  |
| Other subgroup, please specify below |  |  |  |  |  |  |

Comment:

#### 27. About your patients

##### 26. Please indicate the frequency that you include discussion about the following topics with your patients (including new and follow-up). (Please select one response for each topic)

|  | Daily | Weekly | Monthly | Quarterly | Yearly | Never |
| --- | --- | --- | --- | --- | --- | --- |
| Diet or nutrition |  |  |  |  |  |  |
| Smoking, drugs or alcohol |  |  |  |  |  |  |
| General physical activity (not specific MSK exercise rehab) |  |  |  |  |  |  |
| Occupational health and safety or ergonomics |  |  |  |  |  |  |
| Stress management |  |  |  |  |  |  |
| Nutritional supplements (including vitamins, minerals, herbs) |  |  |  |  |  |  |
| Medication (including for pain/inflammation) |  |  |  |  |  |  |
| Infant feeding advice |  |  |  |  |  |  |
| Pain science education |  |  |  |  |  |  |
| Other health promotion advice or education (please specify below) |  |  |  |  |  |  |

Comment:

#### 28. About your patients

##### 27. Please indicate the frequency that you use the following techniques/approaches in your patient management (including new and follow-up). (Please select one response for each technique/approach)

|  | Daily | Weekly | Monthly | Quarterly | Yearly | Never |
| --- | --- | --- | --- | --- | --- | --- |
| Soft tissue massage |  |  |  |  |  |  |
| Muscle Energy Technique (MET) or Proprioceptive Neuromuscular Facilitation (PNF) |  |  |  |  |  |  |
| High Velocity Thrust (HVT) or spinal manipulation/adjustment |  |  |  |  |  |  |
| Spinal articulation or mobilisation |  |  |  |  |  |  |
| Trigger point release |  |  |  |  |  |  |
| General Osteopathic Treatment (GOT) or General Body Adjustment (GBA) |  |  |  |  |  |  |
| Strain-Counterstrain or Facilitated Positional Release |  |  |  |  |  |  |
| Exercise recommendation |  |  |  |  |  |  |
| Cranial osteopathy |  |  |  |  |  |  |
| Neurodynamics or flossing |  |  |  |  |  |  |
| Instrument assisted soft-tissue (e.g. Graston) |  |  |  |  |  |  |
| Taping |  |  |  |  |  |  |
| Kinesiotaping |  |  |  |  |  |  |
| Visceral |  |  |  |  |  |  |
| Dry needling or acupuncture |  |  |  |  |  |  |
| Lymphatic/drainage |  |  |  |  |  |  |
| Extracorporeal Shockwave Therapy (ESWT) |  |  |  |  |  |  |
| Ultrasound |  |  |  |  |  |  |
| Transcutaneous Electrical Nerve Stimulation (TENS) or Electrical Muscle Stimulation (EMS) |  |  |  |  |  |  |
| Intervertebral Differential Dynamics (IDD) therapy or Intermittent Sustained Spinal Traction (ISST) |  |  |  |  |  |  |
| Laser therapy |  |  |  |  |  |  |
| Other technique or approach, please specify below |  |  |  |  |  |  |

Comment:

#### 29. About you

##### 28. How would you describe your gender? (Please select one option only)

|  | Man (including transgender man) |
| --- | --- |
|  | Woman (including transgender woman) |
|  | Prefer not to say |
|  | Prefer to self-describe (e.g., non-binary, gender-fluid, agender), please specify below |

Comment:

#### 30. About you

##### 29. What is your age? (Please select one option only)

|  | 20 – 29 years |
| --- | --- |
|  | 30 – 39 years |
|  | 40 – 49 years |
|  | 50 – 59 years |
|  | 60 – 69 years |
|  | 70 – 79 years |
|  | 80 – 89 years |
|  | 90 years or over |
|  | Prefer not to say |

#### 31. About you

##### 30. How would you describe your ethnicity? (Please select one option only)

|  | White English, Welsh, Scottish, Northern Irish or British |
| --- | --- |
|  | Indian |
|  | Pakistani |
|  | Bangladeshi |
|  | Chinese |
|  | Any other Asian background |
|  | African |
|  | Caribbean |
|  | Any other Black, African or Caribbean background |
|  | White and Black Caribbean |
|  | White and Black African |
|  | White and Asian |
|  | Any other Mixed or Multiple ethnic background |
|  | White Irish |
|  | Gypsy or Irish Traveller |
|  | Any other White background |
|  | Arab |
|  | Any other ethnic group |
|  | Prefer not to say |

#### 32. About you

##### 31. How long have you been an osteopath? (Please enter number of years)

#### 33. About you

##### 32. Please indicate all your qualifications related to healthcare. (Please select all that apply)

|  | Diploma (E.g., DO) |
| --- | --- |
|  | Bachelor’s degree (E.g., BSc or BOst) |
|  | Post-graduate Certificate (PgCert) |
|  | Post-graduate Diploma (PgDip) |
|  | Under-graduate Masters (E.g., MOst) |
|  | Post-graduate Masters (MSc or MA) |
|  | Master of Research (MRes) |
|  | Master of Philosophy (MPhil) |
|  | Professional Doctorate (ProfDoc) |
|  | Philosophy Doctorate (PhD) |

#### 34. About you

##### 33. Do you belong to any professional groups? (Please select all that apply)

No

NCOR Regional Hub

Regional Osteopathic CPD group

Institute of Osteopathy

Osteopathic Sports Care Association

Osteopathy Performing Arts Care Association

Sutherland Cranial College of Osteopathy

Institute of Classical Osteopathy

Other (please specify):

#### 35. About you

##### 34. To what extent do you agree with the following statements? (Please select one response for each statement)

|  | Strongly Agree | Agree | Disagree | Strongly Disagree | Not Sure |
| --- | --- | --- | --- | --- | --- |
| I consider myself to be an evidence-based or evidence-informed osteopath |  |  |  |  |  |
| I regularly read osteopathic peer-reviewed journal publications |  |  |  |  |  |
| I regularly read health-related peer-reviewed journal publications |  |  |  |  |  |
| I currently integrate research findings into my role as a healthcare professional |  |  |  |  |  |
| I currently integrate relevant guidelines into my role as a healthcare professional |  |  |  |  |  |
| I would like to integrate research findings into my role as a healthcare professional more than I currently do |  |  |  |  |  |
| I have research training or experience beyond my undergraduate training as an osteopath |  |  |  |  |  |
| I am interested in being involved in research related to osteopathic practice |  |  |  |  |  |
| I have some spare time to devote to taking part in research related to osteopathic practice |  |  |  |  |  |
| I feel I have the research skills necessary to take an active role in research related to osteopathic practice |  |  |  |  |  |
| I feel I have the research experience necessary to take an active role in research related to osteopathic practice |  |  |  |  |  |
| I would expect to be paid for participating in any research activities |  |  |  |  |  |

#### 36. About you

##### 35. What is your preferred way to receive training related to your role as an osteopath (e.g., CPD activities)? (Please select one option only)

|  | Face to face small group seminars or workshops with other osteopaths only |
| --- | --- |
|  | Face to face small group seminars or workshops with osteopaths and other healthcare professionals |
|  | Face to face osteopathy conferences |
|  | Face to face health-related conferences |
|  | Live (i.e., conducted in real time) online webinars or workshops where you are able to interact with the presenter and other attendees |
|  | Live (i.e., conducted in real time) online presentations where you are NOT able to interact with the presenter and other attendees |
|  | Pre-recorded presentations that can be viewed at any time online |
|  | Structured activities accessible online (not including live presentations) that require you to undertake guided tasks |

#### 37. About you

##### 36. While the activities of the NCOR Research Network will be coordinated by NCOR, we may require regional coordinators or ‘Research Champions’ to oversee research activity at a local level.  These positions will be of interest to osteopaths who would like to undertake further training and gain experience in research and have more responsibility and involvement with the NCOR Research Network.  It is possible that these roles will be paid.  Please indicate whether this is something you may be interested in.

|  | Yes |
| --- | --- |
|  | No |
|  | Maybe |

#### 38. Contact details

##### 37. Please enter you first name

##### 38. Please enter your last name

##### 39. Please enter your email address
